# Estimating Impact of Austerity policies in COVID-19 fatality rates: Examining the dynamics of economic policy and Case Fatality Rates (CFR) of COVID-19 in OECD countries

**DOI:** 10.1101/2020.04.03.20047530

**Authors:** Dawa Sherpa

## Abstract

The paper will attempt to estimate factors which determine the variability of case fatality rates of COVID-19 across OECD countries in the recent time. The objective of the paper is to estimate the impact of government health policies on fatality rates (Case fatality rates) of COVID-19 in_OECD countries while controlling for other demographic and economic characteristics. The analysis is done using non-parametric regression method, i.e. Quantile regression. The result from quantile regression analysis shows that a policy of Austerity (health expenditure cuts) significantly increases the mortality rates of COVID-19 in OECD countries. The policy implication of the study is the need for a robust public-funded health system with wider accessibility to deal with major public health crisis like COVID-19 pandemic.

## 1. Introduction

The contemporary world is facing an unprecedented public health crisis emerging from Covid-19. Covid-19 has spread to 200 countries and infected 877422 people across the world. Out of the total infected people across the globe, nearly 43537 have died, and 185241 have recovered till 1^st^ April 2020 (CSSE, 2020). After the outbreak of Covid-19 and declaration of it being a Pandemic by WHO, there has been a massive increase in the volume of research on Covid-19 (Heymann&Shindo, 2020; Novel, 2020). However, most of the research is restricted to clinical perspectives including SARS Cov-2 reproduction rate (Liu et al., 2020), fatality ratio (Onder et al., 2020; Wu & McGoogan, 2020), asymptotic transmission mode (Bai et al., 2020) and other epidemiological characteristics (Atkeson, 2020; Lipsitch et al., 2020; Remuzzi & Remuzzi, 2020; Rothan & Byrareddy, 2020; Xu et al., 2020). Countries across the globe have responded with various measures including rapid testing of population, isolating suspected individuals, imposing strict social distancing norms and totally shut down of economic activities in the form of lockdowns (Ebrahim et al., 2020; Kupferschmidt & Cohen, 2020; Tanne et al., 2020; Wong et al., 2020). The economic impact of Covid-19 for different regions and countries are studied using different economic models and estimation technique (Abiad et al., 2020; Atkeson, 2020; Fernandes, 2020; Hartley & Makridis, 2020; McKibbin & Fernando, 2020; Ruiz Estrada, 2020).

### 2. Objective of Research

Review of existing literature on Covid-19 manifests a lacuna with respect to the dynamic interplay between Covid-19 and country-specific health policies. This paper attempts to fill this gap by highlighting the interrelationship between long term structural health policies and the Covid-19 fatality rates among Organisation for Economic Co-operation and Development (OECD) countries ^**2**^. Austerity policy is defined as a widespread cut on government expenditure which is targeted to reduce government fiscal deficit and enhance economic growth (Konzelmann, 2014;). Such a significant reduction in government spending has a disproportionately negative impact on government social sector expenditure (Health, Education, Social security etc.). The negative impact of austerity policies in terms of lowering employment, economic growth and increasing inequality is well studied (Blyth, 2013; Krugman, 2015; Stiglitz, 2012; UNCTAD, 2017). In the post-2008 crisis period and under the impact of rising debts burdens, many European countries imposed policies of austerity in 2010. The most severe austerity policies were implemented in Greece, Hungary, Ireland, Latvia, Spain and Portugal (Leschke et al., 2015). the OECD group, there was variation in the extent of reduction in their health expenditure across countries in pursuit of Austerity policies (fiscal consolidation) (Van Gool& Pearson, 2014). The negative impact of such drastic fund cuts on access to health facilities and health indicators is well documented in many OECD countries (Antonakakis& Collins, 2014; Ayuso-Mateos et al., 2013; Ifanti et al., 2013; Kentikelenis et al., 2014, 2014, 2014; Loopstra et al., 2016, 2016; McKee et al., 2012, 2012; Reeves et al., 2014, 2014; Ruckert&Labonté, 2017; Stuckler et al., 2017). So under this background of such drastic cuts in health expenditure, this paper will evaluate the impact of austerity policies (health expenditure cuts) on fatality rates of Covid-19 after controlling for other socio-demographic characteristics which have a significant impact on fatality rates of Covid-19. The fatality rates are measured by crude Case Fatality Rates (CFC), which is the ratio of confirmed death to confirmed positive cases of covid-19 for each country.

### 3. Data source and Methodology

Data used for analysis is taken from various sources. Table 1 provides the list of variables used in the analysis with their respective data sources along with the nature of data.

**Table 1:**
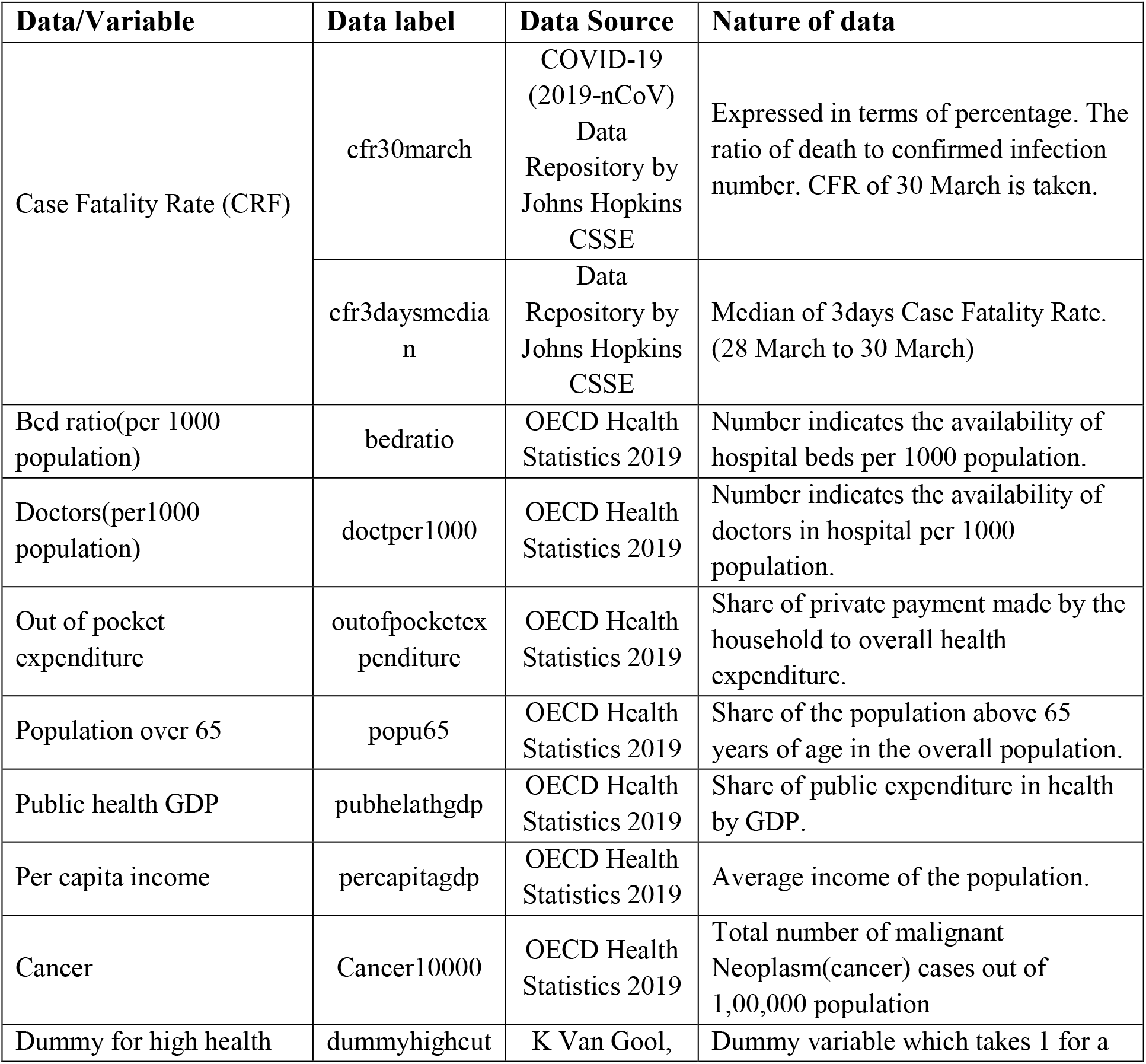
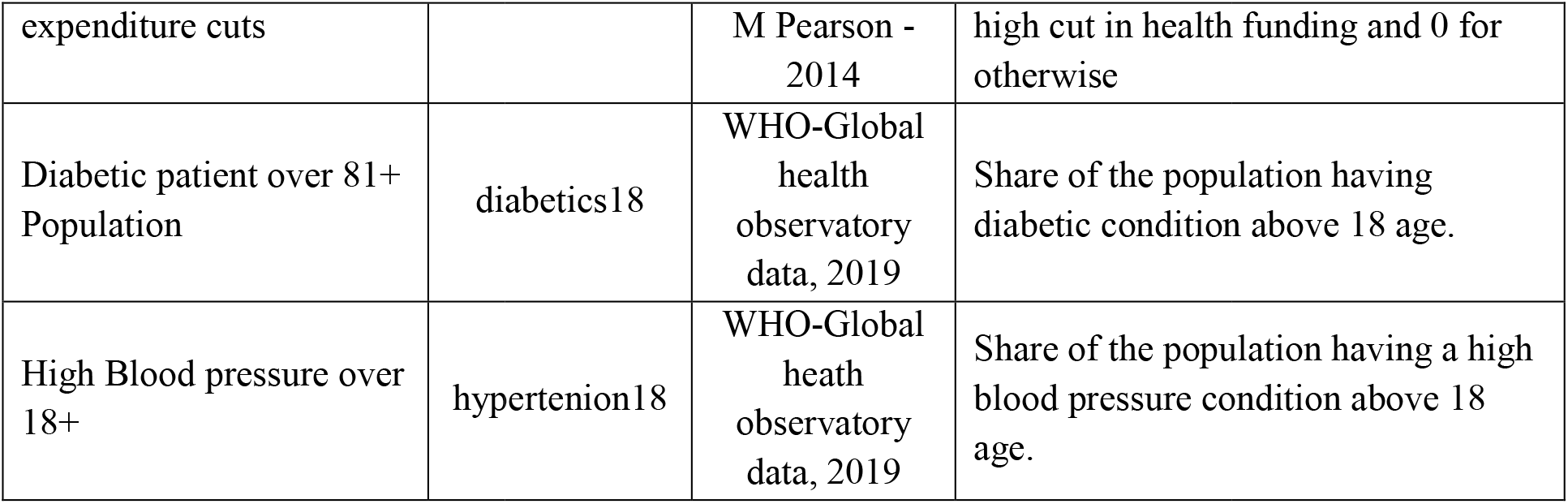
Data labelling and Data source.

The analysis is done using data for thirty-six countries^3^from the OECD group.

Figure1 (a, b) displays that a large part of infection and deaths cases from Covid-19 in the world is concentrated in OECD countries.

**Figure 1:**
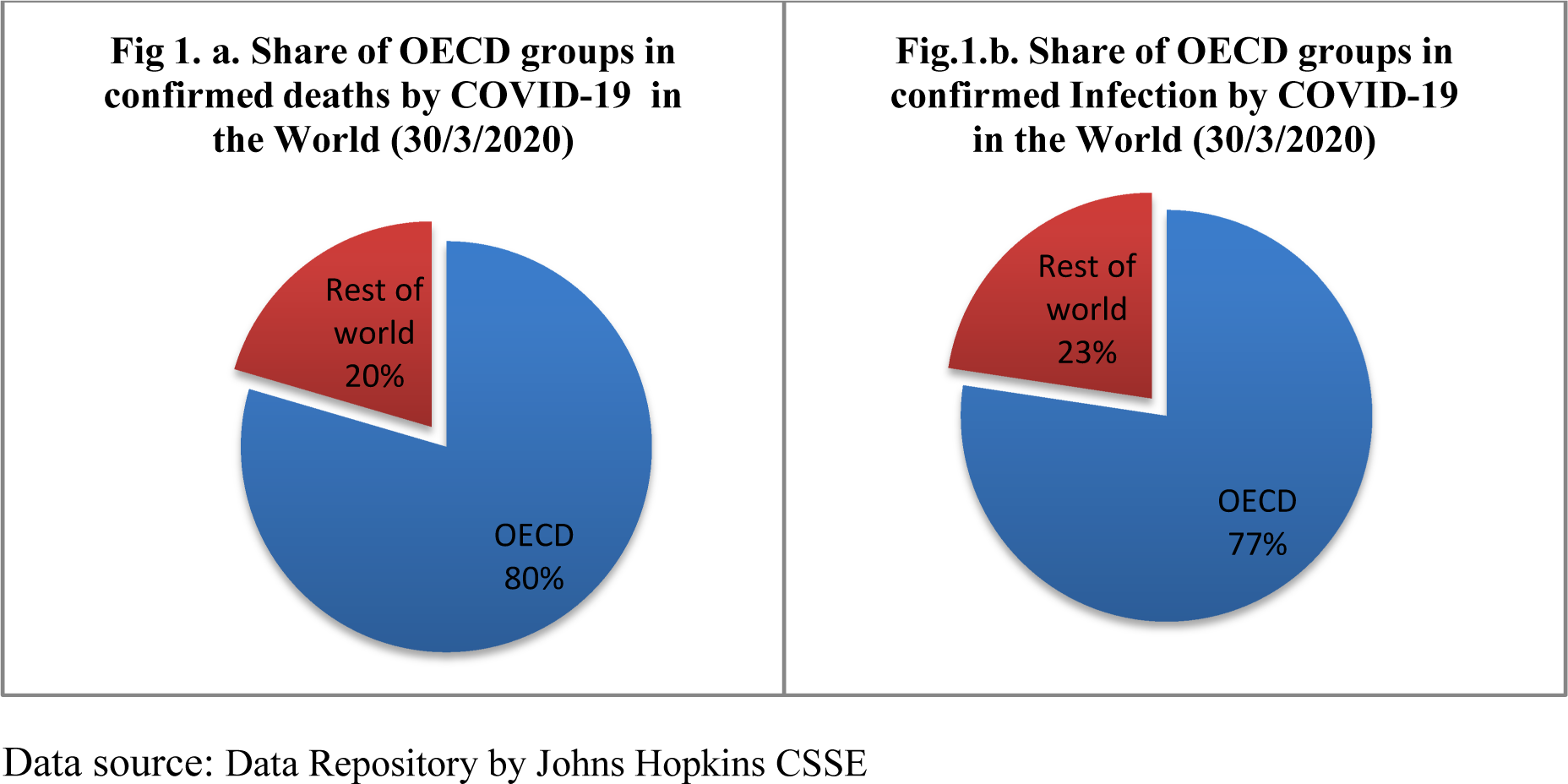
Distribution of Infection and Deaths from COVID-19.

The analysis of the impact of austerity on Covid-19 fatality rates is done using Quantile regression after controlling for all other socio-demographic characteristics which have an impact on Case Fatality Rate (Novel, 2020; Onder et al., 2020; Porcheddu et al., 2020; Wu & McGoogan, 2020). The advantage of using Quantile regression over normal Ordinary Least Square regression (OLS)regression is that it provides a more detailed picture of the relationship between variables not only around mean values but across the distribution of variables (Koenker&Hallock, 2001). It is distribution-free, robust to outliers and capable of modelling entire conditional distribution (Baum, 2013; Cade & Noon, 2003; Yu et al., 2003).

### 4. Statistical Analysis

Table 2 shows the descriptive statistics of all the variables. The mean value of Case Fatality Rate (of 30 March 2020) is 2.554 with a standard deviation of 2.79. The highest value taken by Case Fatality Rate (cfr30march) is 11.6, whereas the smallest value is 0. Similarly, the other variable for Case Fatality Rate, which is cfr3daymedian, also has a similar kind of mean and standard deviation as the previous cfr30march variable.

**Table 2:** Descriptive statistics of variables.

**a. Table 2: Descriptive statistics of variables**
| Variables | N | Mean | Standard deviation | Min | Max | Skewness | Kurtosis |
| --- | --- | --- | --- | --- | --- | --- | --- |
| cfr30march | 36 | 2.554 | 2.792 | 0 | 11.66 | 1.698 | 5.264 |
| cfr3daysmedian | 36 | 2.499 | 2.761 | 0 | 11.59 | 1.676 | 5.276 |
| outofpocketexpenditure | 36 | 22.79 | 12.33 | 7.8 | 65.2 | 1.379 | 5.048 |
| pubhelathgdp | 36 | 5.956 | 2.733 | 0.9 | 14.4 | 0.63 | 3.844 |
| cancer100000 | 36 | 249.1 | 62.01 | 94 | 338.1 | -0.718 | 2.674 |
| peccapitagdp | 36 | 34,597 | 24,264 | 1,923 | 104,499 | 0.684 | 3.137 |
| doctper1000 | 36 | 3.119 | 1.002 | 0.32 | 5.18 | -0.574 | 3.843 |
| dummyhighcut | 36 | 0.389 | 0.494 | 0 | 1 | 0.456 | 1.208 |
| hypertenion18 | 36 | 26.08 | 6.315 | 13.2 | 38.2 | 0.132 | 2.273 |
| diabetics18 | 36 | 8.372 | 1.674 | 5.6 | 13.2 | 0.514 | 3.236 |
| popu65 | 36 | 15.94 | 5.457 | 5.4 | 28.2 | -0.338 | 2.552 |
| bedratio | 36 | 4.292 | 2.785 | 0.53 | 13.05 | 1.554 | 5.382 |

The average value of public spending in health to GDP variable, publichelathgdp, is 5.95 % and it has a minimum value of 0.9 per cent and the maximum value of 14.4 per cent. The existing clinical research shows that the fatality rate of Covid-19 is influenced by the existence of pre-medical complication and the share of older adults in the population (Onder et al., 2020; Wu & McGoogan, 2020). The crucial demographic variable, population share above 65 years (popu65), has a mean value of 15 per cent and a standard deviation of 5.4. The share of the population having hypertension (above 18 years of age) has a minimum value of 13 % and a maximum value of 38.2 %.

#### b. Quantile Regression

Table 3 shows the result of Quantile regression. The first model has Case Fatality Rate of March 30 as the dependent variable. In the second model, the dependent variable is the three-day median Case Fatality Rate. The results from both models show that the coefficient of the dummy variable for high fund cut has a positive impact on CFR and is significant at one per cent level of significance. The results show that a country which has a history of drastic health fund cut is increasing the fatality rates from Covid-19. Similarly, the coefficient of the variable of public health GDP is negative and significant at one per cent level of significance which implies that countries which have a higher share of public-funded health system have lower Case Fatality Rates.

**Table 3:** Result of the Quantile Regression.

|  | (Model -1) | (Model-2) |
| --- | --- | --- |
| **VARIABLES** | **cfr30march** | **cfr3dysmedian** |
| dummyhighcut | 0.660***<br>(0.171) | 0.798***<br>(0.159) |
| cancer100000 | -0.00701***<br>(0.00229) | -0.00644***<br>(0.00220) |
| hypertenion18 | 0.0541***<br>(0.0128) | 0.0511***<br>(0.0125) |
| diabetics18 | -0.100**<br>(0.0409) | -0.102**<br>(0.0406) |
| pubhelathgdp | -0.344***<br>(0.0368) | -0.361***<br>(0.0331) |
| doctper1000 | -0.366***<br>(0.0892) | -0.411***<br>(0.0722) |
| bedratio | -0.0491<br>(0.0340) | -0.0668**<br>(0.0315) |
| outofpocketexpenditure | -0.00832<br>(0.00769) | 0.000757<br>(0.00737) |
| popu65 | 0.107***<br>(0.0215) | 0.120***<br>(0.0194) |
| peccapitagdp | 1.59e-05***<br>(4.91e-06) | 1.62e-05***<br>(4.54e-06) |
| Constant | 4.190***<br>(0.635) | 4.084***<br>(0.639) |
| Observations | 36 | 36 |

Standard errors in parentheses \*\*\* p<0.01, \*\* p<0.05, \* p<0.1

The impact of good health infrastructure (measured by bed per 1000 population and doctor per 1000 population) on Case Fatality Rates is negative. Anticipated as per the existing literature, the result confirms that countries having higher share of old age population have higher fatality rates. Also, higher the share of pre-existing medical condition in the overall population, higher is the fatality rate from COVID-19.

The presence of model specification error is done using link test. If the regression model does not contain specification error, then the variable **_hatsq** will be statistically insignificant. **Table 4** shows the result of the link test for Model −1. The P-value of the variable **_hatsq** is 0.16, and hence it is statistically insignificant. Therefore it can be concluded thatModel-1 does not contain specification error.

**Table 4:** Model Specification Test: Link Test of Model-1.

|  |  |  |
| --- | --- | --- |
| Median regression | Number of obs = | 36 |
| Raw sum of deviations 65.76895 (about 1.6081871) |  |  |
| Min sum of deviations 56.4424 | Pseudo R2 = | 0.1418 |

| cfr30march | Coef. | Std. Err. | t | P>\|t\| | [95% Conf. Interval] |  |
| --- | --- | --- | --- | --- | --- | --- |
| _hat | .9760163 | .0240544 | 40.58 | 0.000 | .9270773 | 1.024955 |
| _hatsq | .0088423 | .0062851 | 1.41 | 0.169 | -.0039449 | .0216295 |
| _cons | .0147506 | .0207319 | 0.71 | 0.482 | -.0274287 | .0569299 |

**Table 5:** Model Specification Test: Link Test of Model-2.

|  |  |  |
| --- | --- | --- |
| Median regression | Number of obs = | 36 |
| Raw sum of deviations 65.56853 (about 1.6014235) |  |  |
| Min sum of deviations 56.50638 | Pseudo R2 = | 0.1382 |

| cfr3daysme~n | Coef. | Std. Err. | t | P>\|t\| | [95% Conf. Interval] |  |
| --- | --- | --- | --- | --- | --- | --- |
| _hat | 1 | .0273136 | 36.61 | 0.000 | .94443 | 1.05557 |
| _hatsq | 1.20e-09 | .0068069 | 0.00 | 1.000 | -.0138488 | .0138488 |
| _cons | 5.75e-09 | .0247473 | 0.00 | 1.000 | -.0503488 | .0503488 |

Similarly, the link test result of Model-2 also shows that the variable **_hatsq** is not statistically significant. Hence Model-2 also does not contain specification error.

In order to obtain an idea about the coefficients of Quantile regression of independent variables across quantiles of Case Fatality Rates, following figures (2 and 3) have been derived using the Azevedo method (Azevedo, 2011). It shows how the impact of each independent variable varies across quantiles.

Figure 2 and 3 show that the coefficients of Dummy variable (for health fund cut) of the Quantile regression is positive and increases across quantiles of Case Fatality Rate Only for the third quantile, the coefficient is negative. So the impact of Austerity is positive on Case Fatality Rate of Covid-19 except for the third quantile.

**Figure 2:**
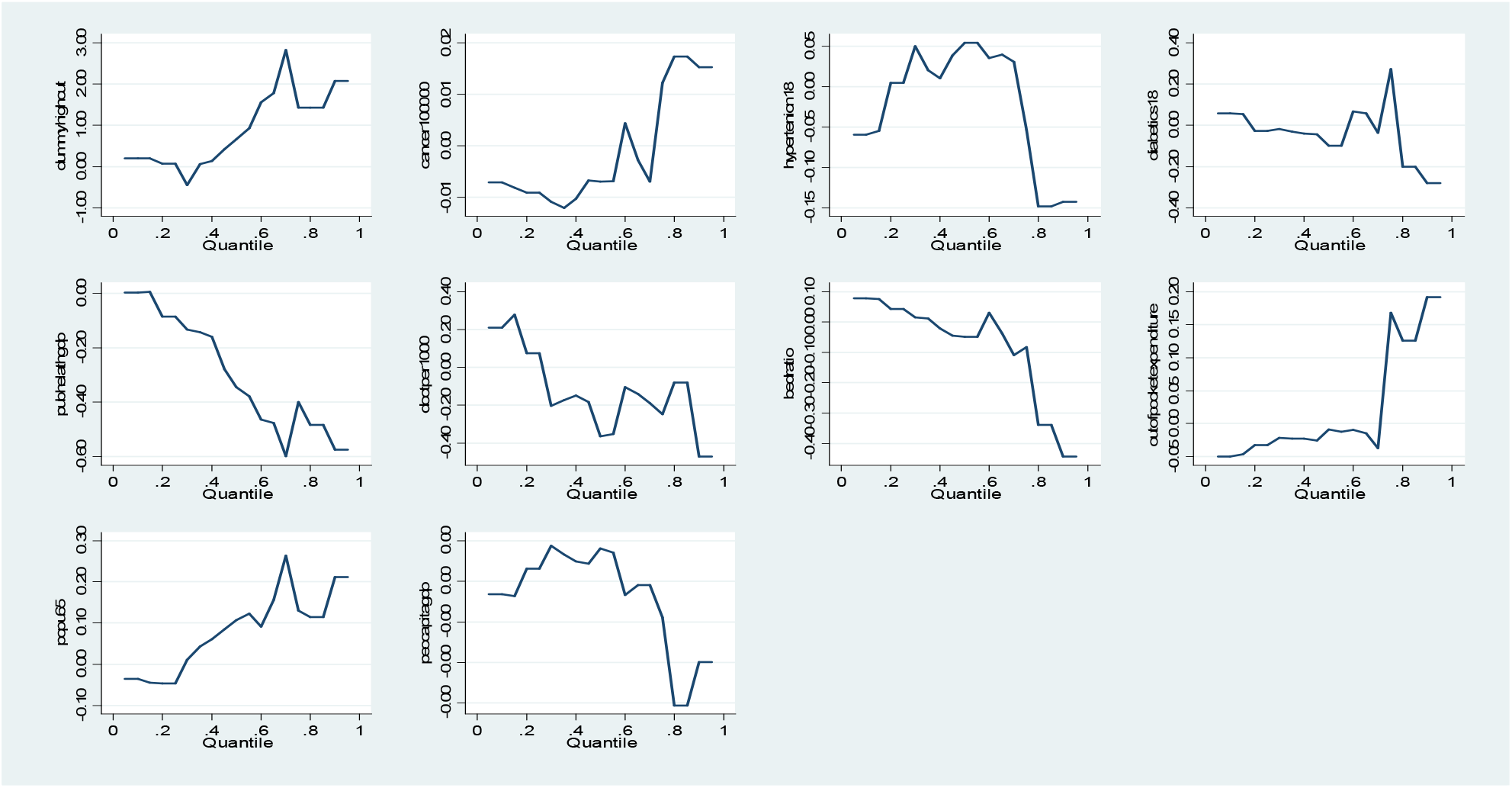
The coefficients of a Quantile Regression (Model-1)

**Figure 3:**
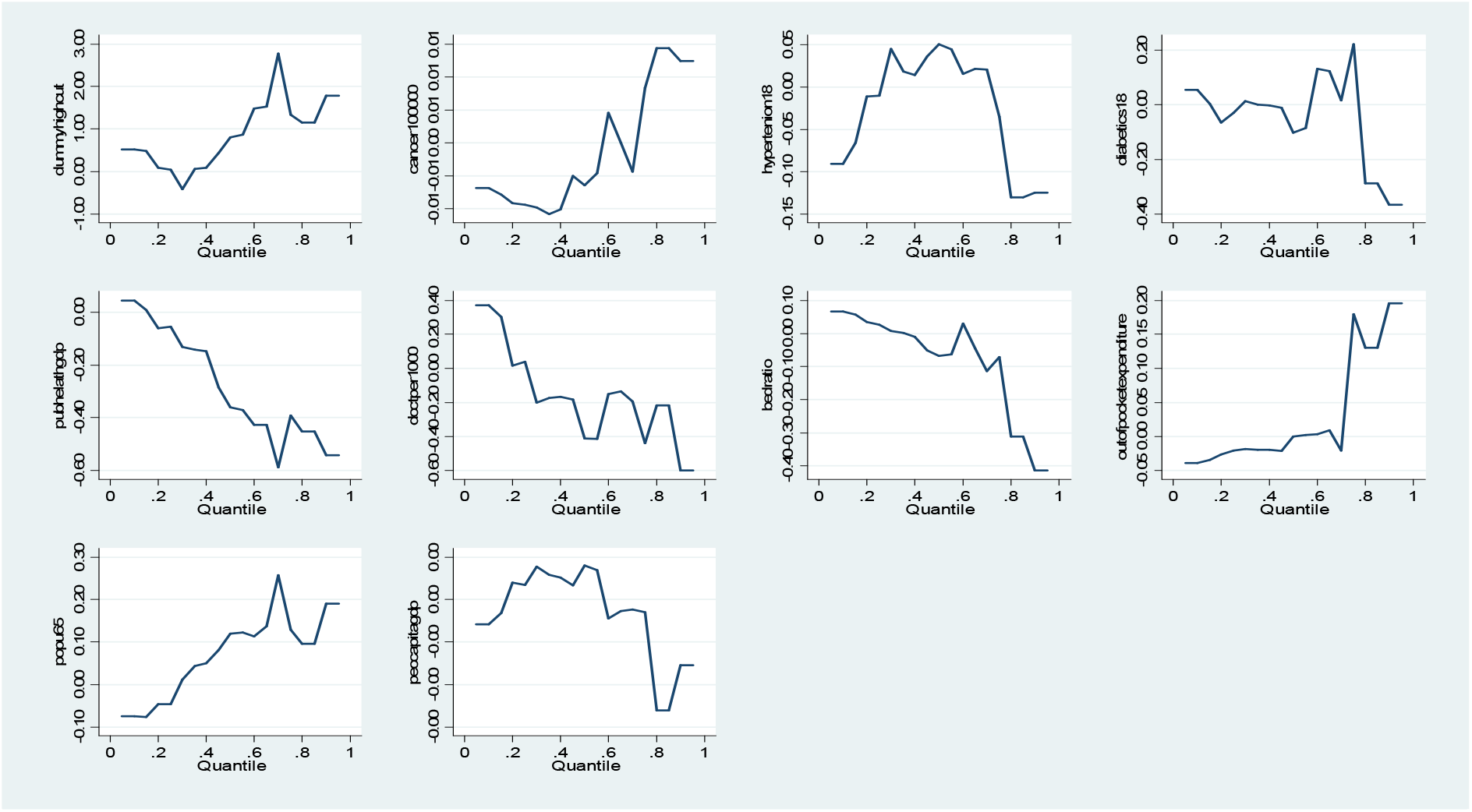
The coefficients of a Quantile Regression (Model-2)

The coefficients of the public fund on health to GDP variable is negative (except for the first quantile) across the distribution of Case Fatality Rate It indicates that higher public expenditure on health reduces fatality rates of Covid-19. The coefficients of doctor per 1000 population is negative across the distribution of Case Fatality Rate (except for the second quantile). The coefficients of hospital bed per 1000 population is negative across the distribution of Case Fatality Rate (except till the fourth quantile).

## 5. Conclusion

The result from the Quantile regression analysis shows that countries which have pursued austerity policies has significantly higher fatality rates from COVID-19 after controlling for all other socio-demographic factors which influence Case Fatality Rate of COVID-19. Higher public funding share, higher doctors per population, higher bed availability is associated with lower fatality rates from COVID-19. A higher share of population with pre-medical conditions (diabetics, hypertension) and older age population increase fatality rates. So the policies of austerity (at least in terms of reduction in health expenditure) can significantly worsen the health system’s ability to fight pandemic like COVID-19 and can lead to a severe negative health outcome. The policy implication of the study is the need for a robust public-funded health system with wider accessibility to deal with major public health crisis like theCovid-19 pandemic.

## Data Availability

Research is based on secondary data which is publicly available.

https://www.oecd.org/health/health-data.htm

https://www.who.int/data/gho/data/indicators

## Footnotes

2 Member countries of OECD group are Australia, Austria, Belgium, Canada, the Czech Republic, Denmark, Finland, France, Germany, Greece, Hungary, Iceland, Ireland, Italy, Japan, Korea, Luxembourg, Mexico, The Netherlands, New Zealand, Norway, Poland, Portugal, the Slovak Republic, Spain, Sweden, Switzerland, Turkey, the United Kingdom, and the United States.

## References

Abiad, A., Arao, R. M., & Dagli, S. (2020). The Economic Impact of the COVID-19 Outbreak on Developing Asia.

Antonakakis, N., & Collins, A. (2014). The impact of fiscal austerity on suicide: On the empirics of a modern Greek tragedy. Social Science & Medicine, 112, 39–50.

Atkeson, A. (2020). What Will Be the Economic Impact of COVID-19 in the US? Rough Estimates of Disease Scenarios. National Bureau of Economic Research.

Ayuso-Mateos, J. L., Barros, P. P., & Gusmão, R. (2013). Financial crisis, austerity, and health in Europe. The Lancet, 382(9890), 391–392.

Azevedo, J. P. (2011). grqreg: Stata module to graph the coefficients of a quantile regression.

Bai, Y., Yao, L., Wei, T., Tian, F., Jin, D.-Y., Chen, L., & Wang, M. (2020). Presumed asymptomatic carrier transmission of COVID-19. Jama.

Baum, C. F. (2013). Quantile regression. URL http://Fmwww.Bc.Edu/EC-CS, 2013.

Bénassy-Quéré, A., Marimon, R., Pisani-Ferry, J., Reichlin, L., Schoenmaker, D., & Weder, B. (2020). 13 COVID-19: Europe needs a catastrophe relief plan. Mitigating the COVID Economic Crisis: Act Fast and Do Whatever, 121.

Blyth, M. (2013). Austerity: The history of a dangerous idea. Oxford University Press.

Cade, B. S., & Noon, B. R. (2003). A gentle introduction to quantile regression for ecologists. Frontiers in Ecology and the Environment, 1(8), 412–420.

Csse, J. (2020). Coronavirus COVID-19 Global Cases by the Center for Systems Science and Engineering (CSSE) at Johns Hopkins University (JHU). In 2020-03-15]. Https://gisanddata.Maps.Arcgis.Com/apps/opsdashboard/index.Html#/bda7594740fd40299423467b48e9ecf6.

del Rio, C., & Malani, P. N. (2020). COVID-19—New insights on a rapidly changing epidemic. Jama.

Ebrahim, S. H., Ahmed, Q. A., Gozzer, E., Schlagenhauf, P., & Memish, Z. A. (2020). Covid-19 and community mitigation strategies in a pandemic. British Medical Journal Publishing Group.

Fernandes, N. (2020). Economic effects of coronavirus outbreak (COVID-19) on the world economy. Available at SSRN 3557504.

Hartley, J., & Makridis, C. (2020). Forecasting County-level Real GDP Effects of COVID-19. Available at SSRN 3559139.

Heymann, D. L., & Shindo, N. (2020). COVID-19: What is next for public health? The Lancet, 395(10224), 542–545.

Ifanti, A. A., Argyriou, A. A., Kalofonou, F. H., & Kalofonos, H. P. (2013). Financial crisis and austerity measures in Greece: Their impact on health promotion policies and public health care. Health Policy, 113(1–2), 8–12.

Kentikelenis, A., Karanikolos, M., Reeves, A., McKee, M., & Stuckler, D. (2014). Greece’s health crisis: From austerity to denialism. The Lancet, 383(9918), 748–753.

Koenker, R., & Hallock, K. F. (2001). Quantile regression. Journal of Economic Perspectives, 15(4), 143–156.

Konzelmann, S. J. (2014). The political economics of austerity. Cambridge Journal of Economics, 38(4), 701–741.

Krugman, P. (2015). The austerity delusion. The Guardian, 29, 31–3.

Kupferschmidt, K., & Cohen, J. (2020). Can China’s COVID-19 strategy work elsewhere? American Association for the Advancement of Science.

Leschke, J., Theodoropoulou, S., & Watt, A. (2015). Towards ‘Europe 2020’? Austerity and new economic governance in the EU. Edited By, 295.

Lipsitch, M., Swerdlow, D. L., & Finelli, L. (2020). Defining the epidemiology of Covid-19—Studies needed. New England Journal of Medicine.

Liu, Y., Gayle, A. A., Wilder-Smith, A., & Rocklöv, J. (2020). The reproductive number of COVID-19 is higher compared to SARS coronavirus. Journal of Travel Medicine.

Loopstra, R., McKee, M., Katikireddi, S. V., Taylor-Robinson, D., Barr, B., & Stuckler, D. (2016). Austerity and old-age mortality in England: A longitudinal cross-local area analysis, 2007–2013. Journal of the Royal Society of Medicine, 109(3), 109–116.

McKee, M., Karanikolos, M., Belcher, P., & Stuckler, D. (2012). Austerity: A failed experiment on the people of Europe. Clinical Medicine, 12(4), 346.

McKibbin, W., & Fernando, R. (n.d.). 3 The economic impact of COVID-19. Economics in the Time of COVID-19, 45.

McKibbin, W. J., & Fernando, R. (2020). The global macroeconomic impacts of COVID-19: Seven scenarios.

Novel, C. P. E. R. E. (2020). The epidemiological characteristics of an outbreak of 2019 novel coronavirus diseases (COVID-19) in China. Zhonghua Liu Xing Bing Xue Za Zhi= Zhonghua Liuxingbingxue Zazhi, 41(2), 145.

Onder, G., Rezza, G., & Brusaferro, S. (2020). Case-fatality rate and characteristics of patients dying in relation to COVID-19 in Italy. JAMA.

Porcheddu, R., Serra, C., Kelvin, D., Kelvin, N., & Rubino, S. (2020). Similarity in Case Fatality Rates (CFR) of COVID-19/SARS-COV-2 in Italy and China. The Journal of Infection in Developing Countries, 14(02), 125–128.

Reeves, A., McKee, M., Basu, S., & Stuckler, D. (2014). The political economy of austerity and healthcare: Cross-national analysis of expenditure changes in 27 European nations 1995– 2011. Health Policy, 115(1), 1–8.

Remuzzi, A., & Remuzzi, G. (2020). COVID-19 and Italy: What next? The Lancet.

Rothan, H. A., & Byrareddy, S. N. (2020). The epidemiology and pathogenesis of coronavirus disease (COVID-19) outbreak. Journal of Autoimmunity, 102433.

Ruckert, A., & Labonté, R. (2017). Health inequities in the age of austerity: The need for social protection policies. Social Science & Medicine, 187, 306–311.

Ruiz Estrada, M. A. (2020). Economic Waves: The Effect of the Wuhan COVID-19 On the World Economy (2019-2020). Available at SSRN 3545758.

Stiglitz, J. (2012). Austerity–Europe’s man-made disaster. Social Europe Journal, 8.

Stuckler, D., Reeves, A., Loopstra, R., Karanikolos, M., & McKee, M. (2017). Austerity and health: The impact in the UK and Europe. European Journal of Public Health, 27(suppl_4), 18–21.

Tanne, J. H., Hayasaki, E., Zastrow, M., Pulla, P., Smith, P., & Rada, A. G. (2020). Covid-19: How doctors and healthcare systems are tackling coronavirus worldwide. Bmj, 368.

UNCTAD. (2017). Trade and Development Report 2017–Beyond austerity: Towards a global New Deal. United Nations New York and Geneva.

Van Gool, K., & Pearson, M. (2014). Health, austerity and economic crisis.

Wang, C. J., Ng, C. Y., & Brook, R. H. (2020). Response to COVID-19 in Taiwan: Big data analytics, new technology, and proactive testing. JAMA.

Wong, J. E., Leo, Y. S., & Tan, C. C. (2020). COVID-19 in Singapore—current experience: Critical global issues that require attention and action. JAMA.

Wu, Z., & McGoogan, J. M. (2020). Characteristics of and important lessons from the coronavirus disease 2019 (COVID-19) outbreak in China: Summary of a report of 72 314 cases from the Chinese Center for Disease Control and Prevention. Jama.

Xu, B., Kraemer, M. U., Gutierrez, B., Mekaru, S., Sewalk, K., Loskill, A., Wang, L., Cohn, E., Hill, S., & Zarebski, A. (2020). Open access epidemiological data from the COVID-19 outbreak. The Lancet Infectious Diseases.

You, C., Deng, Y., Hu, W., Sun, J., Lin, Q., Zhou, F., Pang, C. H., Zhang, Y., Chen, Z., & Zhou, X.-H. (2020). Estimation of the time-varying reproduction number of COVID-19 outbreak in China. Available at SSRN 3539694.

Yu, K., Lu, Z., & Stander, J. (2003). Quantile regression: Applications and current research areas. Journal of the Royal Statistical Society: Series D (The Statistician), 52(3), 331–350.

